## Additional file 2 for "Implementation strategies and outcomes of intravenous iron use for treatment of anaemia during and after pregnancy in LMICs: a scoping review"

|  | **Intravenous iron** |  |
| --- | --- | --- |
| #1 | “intravenous iron” OR “iv iron” OR “iron infusion” OR “parenteral iron” OR “iron dextran” OR “iron sucrose” OR “sodium ferric gluconate” OR ferumoxytol OR “ferric carboxymaltose” OR “iron isomaltoside” OR “iron polymaltose” OR “iron saccharate” OR “iron carbohydrate” OR “ferric compound*” OR “ferrous compound*” | Title/ Abstract |
| #2 | (MH "Iron+" OR MH "Ferric Oxide, Saccharated" OR MH "Iron-Dextran Complex" OR MH "Ferrosoferric Oxide" OR MH "Ferric Compounds+" OR (MH "Iron Compounds+") | MeSH Terms |
| **#3** | **#1 OR #2** | **169,086** |
|  | **Pregnancy** |  |
| #1 | pregnan* OR expectant OR maternal OR prenatal* OR antenatal* OR gestation* OR obstetric* OR perinatal* OR puerperium OR parturition OR childbearing OR mother* | Title/ Abstract |
| #2 | (MH "Pregnancy" OR MH "Expectant Mothers" OR MH "Prenatal Care" OR MH "Perinatal Care" OR MH "Perinatal Period" OR MH “Obstetric" OR MH "Obstetric Care") | MeSH Terms |
|  | **Postpartum** |  |
| #3 | Postpartum OR postnatal* | Title/ Abstract |
| #4 | (MH “Postpartum Period" OR MH "Postnatal Care") | MeSH Terms |
| **#5** | **#1 OR #2 OR #3 OR #4** | **1,910,234** |
|  | **Low to Middle Income Countries*** |  |
| #1 | MH “Developing Countries” | MeSH Terms |
| #2 | Africa or Asia or Caribbean or West Indies or South America or Latin America or Central America | No fields selected |
| #3 | Afghanistan or Albania or Algeria or Angola or Antigua or Barbuda or Argentina or Armenia or Armenian or Aruba or Azerbaijan or Bahrain or Bangladesh or Barbados or Benin or Byelarus or Byelorussian or Belarus or Belorussian or Belorussia or Belize or Bhutan or Bolivia or Bosnia or Herzegovina or Hercegovina or Botswana or Brasil or Brazil or Bulgaria or Burkina Faso or Burkina Fasso or Upper Volta or Burundi or Urundi or Cambodia or Khmer Republic or Kampuchea or Cameroon or Cameroons or Cameron or Camerons or Cape Verde or Central African Republic or Chad or Chile or China or Colombia or Comoros or Comoro Islands or Comores or Mayotte or Congo or Zaire or Costa Rica or Cote d'Ivoire or Ivory Coast or Croatia or Cuba or Cyprus or Czechoslovakia or Czech Republic or Slovakia or Slovak Republic or Djibouti or French Somaliland or Dominica or Dominican Republic or East Timor or East Timur or Timor Leste or Ecuador or Egypt or United Arab Republic or El Salvador or Eritrea or Estonia or Ethiopia or Fiji or Gabon or Gabonese Republic or Gambia or Gaza or Georgia Republic or Georgian Republic or Ghana or Gold Coast or Greece or Grenada or Guatemala or Guinea or Guam or Guiana or Guyana or Haiti or Honduras or Hungary or India or Maldives or Indonesia or Iran or Iraq or Isle of Man or Jamaica or Jordan or Kazakhstan or Kazakh or Kenya or Kiribati or Korea or Kosovo or Kyrgyzstan or Kirghizia or Kyrgyz Republic or Kirghiz or Kirgizstan or Lao PDR or Laos or Latvia or Lebanon or Lesotho or Basutoland or Liberia or Libya or Lithuania or Macedonia or Madagascar or Malagasy Republic or Malaysia or Malaya or Malay or Sabah or Sarawak or Malawi or Nyasaland or Mali or Malta or Marshall Islands or Mauritania or Mauritius or Agalega Islands or Mexico or Micronesia or Middle East or Moldova or Moldovia or Moldovian or Mongolia or Montenegro or Morocco or Ifni or Mozambique or Myanmar or Myanma or Burma or Namibia or Nepal or Netherlands Antilles or New Caledonia or Nicaragua or Niger or Nigeria or Northern Mariana Islands or Oman or Muscat or Pakistan or Palau or Palestine or Panama or Paraguay or Peru or Philippines or Philipines or Phillipines or Phillippines or Poland or Portugal or Puerto Rico or Romania or Rumania or Roumania or Russia or Russian or Rwanda or Ruanda or Saint Kitts or St Kitts or Nevis or Saint Lucia or St Lucia or Saint Vincent or St Vincent or Grenadines or Samoa or Samoan Islands or Navigator Island or Navigator Islands or Sao Tome or Saudi Arabia or Senegal or Serbia or Montenegro or Seychelles or Sierra Leone or Slovenia or Sri Lanka or Ceylon or Solomon Islands or Somalia or South Africa or Sudan or Suriname or Surinam or Swaziland or Syria or Tajikistan or Tadzhikistan or Tadjikistan or Tadzhik or Tanzania or Thailand or Togo or Togolese Republic or Tonga or Trinidad or Tobago or Tunisia or Turkey or Turkmenistan or Turkmen or Uganda or Ukraine or Uruguay or USSR or Soviet Union or Union of Soviet Socialist Republics or Uzbekistan or Uzbek or Vanuatu or New Hebrides or Venezuela or Vietnam or Viet Nam or West Bank or Yemen or Yugoslavia or Zambia or Zimbabwe or Rhodesia | No fields selected |
| #4 | ((developing or less* developed or under developed or underdeveloped or middle income or low* income or underserved or under served or deprived or poor*) N1 (countr* or nation? or population? or world)) | No fields selected |
| #5 | ((developing or less* developed or under developed or underdeveloped or middle income or low* income) N1 (economy or economies)) | No fields selected |
| #6 | (low* N1 (gdp or gnp or gross domestic or gross national)) | No fields selected |
| #7 | (low N3 middle N3 countr*) | No fields selected |
| #8 | (lmic or lmics or third world or lami countr*) | No fields selected |
| #9 | transitional countr* | No fields selected |
| #10 | ((high burden or high-burden or countdown) N1 countr*) | No fields selected |
| **#11** | **#1 OR #2 OR #3 OR #4 OR #5 OR #6 OR #7 OR #8 OR 9 OR 10** | **8,580,480** |
|  | **#3 AND #5** **AND #11** | **2,811** |
